# Enhancing Early Warning Outbreak Detection Using Multi Model Stacking Ensemble

**DOI:** 10.64898/2026.07.09.26357658

**Authors:** Juliane F. Oliveira, Andrêza L. Alencar, Eluã R. Coutinho, Dérick G. F. Borges, Fábio M. H. S. Filho, Rejane Santos-Silva, Pilar T. V. Florentino, Maria Célia S. L. Cunha, Izabel Marcilio, Roberto F. S. Andrade, Pablo Ivan P. Ramos, Manoel Barral-Netto

## Abstract

**Background:** Evaluating outbreak detection models is a key component of syndromic surveillance. However, balancing timeliness, predictive performance, and local surveillance constraints remains a major challenge. We developed and assessed whether stacking ensemble approaches, which integrate multiple outbreak detection methods, can improve the timeliness and predictive performance of influenza-like illness (ILI) surge detection.

**Methods:** We developed a two-stage stacking ensemble framework to detect early warning of ILI surges in city-level Primary Health Care encounter time series from Brazil (2022–2025). Epidemic thresholds were defined using the Moving Epidemic Method (MEM). In the first stage, multiple outbreak detection models (ODMs) generated warnings of unusual ILI activity. In the second, these warnings were then used as inputs to three supervised meta-classifiers: Logistic Regression, Extreme Gradient Boosting (XGB), and a Multi-layer Perceptron (MLP). For comparison, a Majority Voting (MV) aggregation is also examined. Timeliness, sensitivity, specificity, positive and negative predictive values are evaluated to measure each model’s ability to anticipate epidemic periods of varying intensity in 2025. Robustness was further assessed using simulated outbreak scenarios with varying magnitudes and durations.

**Findings:** We identified 5,765 ILI surge onsets across 5,365 Brazilian municipalities in 2025. Compared with individual ODMs and MV, stacking ensemble meta-classifiers anticipated up to 33% of surge onsets three weeks in advance (an average improvement of 15 percentage points) while reducing missed detections to <10%. They achieved sensitivity >90%, while maintaining balanced specificity >80%, PPV >65%, and NPV >99%. Improvements were greatest for very high-intensity surges, with missed detections reduced by more than half compared with individual ODMs. In simulated outbreak scenarios, the MLP and XGB classifiers remained robust despite being trained on fewer than half of all simulated surge events, consistently outperforming individual detection methods and simpler integration approaches.

**Interpretation:** We provide a practical framework for integrating complementary ODMs into a single, robust early warning decision. By improving both timeliness and predictive performance without requiring additional surveillance data or resources, this approach offers a scalable methodological upgrade for syndromic surveillance systems and supports more reliable public health decision-making.

**Funding:** The Rockefeller Foundation (award 2023 PPI 007 to MB-N); Brazilian National Research Council (CNPq (408775/2024-6); MB-N, PIPR, RFSA are CNPq fellows.

**Research in context:** *Evidence before this study:* We searched PubMed up to June 2025 without language or date restrictions to identify studies evaluating outbreak detection methods (ODMs) for syndromic surveillance (using the terms (“early warning system” OR “syndromic surveillance” OR “outbreak detection“) AND (“infectious disease*” OR “communicable disease*“) AND (“timeliness” OR “sensitivity” OR “specificity” OR “predictive value“)) and studies combining multiple ODMs (using (“outbreak detection model*” OR “aberration detection“) AND (“ensemble” OR “stacking” OR “meta-classifier” OR “model combination“) AND surveillance). Of 458 records screened, 45 were relevant. Most studies (32) compared individual ODMs using simulated outbreaks over synthetic surveillance baselines, reporting sensitivity (approximately 48–99%), specificity, false-alarm rate, and timeliness (hours to a few days). Although ensemble methods are widely used in biomedical machine learning, their application to outbreak detection has been limited. Existing infectious disease applications primarily combine forecasting models or construct reference labels to evaluate early warning systems, rather than integrating first-stage ODM outputs through supervised learning. Only five studies combined multiple ODMs for a single detection decision, and all relied on simple aggregation rules or expert review rather than learned meta-classifiers. The remaining studies (8) are narrative reviews or do not include quantitative data. They particularly point out the absence of standardised evaluation frameworks for syndromic surveillance systems and the need for methods that reflect the heterogeneity of local surveillance contexts and data constraints.

*Added value of this study:* To our knowledge, this is the first study to evaluate a two-stage stacking ensemble in which warnings from multiple independent ODMs serve as input features for supervised meta-classifiers to detect ILI surges using real-world, city-level primary health care data. Unlike prior combination approaches, our framework learns how to weight and integrate first-stage ODM outputs, reports standard surveillance performance metrics, and reflects a representative balance of modelling techniques adaptable to different data constraints and local contexts. We also evaluate the approach using simulated outbreaks superimposed on real municipal surveillance time series, bridging the gap between synthetic benchmarking and operational surveillance.

*Implications of all the available evidence:* Current syndromic surveillance systems based on individual ODMs or simple combination rules often show moderate performances. Our findings suggest that a stacking ensemble framework can meaningfully improve both timeliness and predictive performance of outbreak detection without requiring additional surveillance infrastructure. This offers local health authorities a low-cost methodological upgrade to existing syndromic surveillance pipelines and supports the integration of diverse modelling approaches within routine public health practice.

## Introduction

Syndromic surveillance aims to detect early signs of disease activity before diagnoses are confirmed^1^. A common strategy involves applying outbreak detection models to different streams of time-series data, such as emergency department visits, drug sales, telehealth calls, social media interactions, school absenteeism, primary health care encounters, and other routinely collected data^2^. Use of these conventional and non-conventional data streams enable us to identify outbreaks of infectious diseases earlier than traditional, indicator-based surveillance strategies^2,3^. However, selecting an optimal modeling strategy, or effectively combining multiple approaches, remains difficult. Different outbreak detection models applied to the same data can yield divergent anomaly patterns due to local epidemiological dynamics, model parametrization, and data constraints^4–7^. This model-induced variability can undermine the consistency of outbreak detection, particularly in decentralized surveillance systems where multiple teams may apply different methods independently^8^.

To address the discrepancies, recent studies have explored combining outbreak detection models outputs through direct or indirect integration^9–13^. These approaches generally follow two strategies. The first uses epidemiological indicators, such as weekly healthcare encounters, to train a predictive ensemble of models, often incorporating external data such as web-based, meteorological, demographic or socioeconomic variables. In this ensemble framework, outbreak signals are subsequently derived from forecast errors based on predefined thresholds^10,11^. However, these forecast-based strategies depend on accurate baseline activity estimations, which requires long-term data and may become unstable during epidemiological transitions or under poor data quality^14^.

The second strategy aggregates the binary outputs of multiple outbreak detection models using rules such as majority voting or union criteria to generate a final outbreak warning^9,11,13^. Although promising, these approaches have important limitations. Majority voting assumes equal model reliability, failing to account for context-specific performance differences. In contrast, union rules may increase sensitivity but often at the cost of excessive false alarms, contributing to alert fatigue and reduced trust.

Crucially, many outbreak detection models may lack sufficient anticipatory power and fail to achieve an appropriate balance between sensitivity, specificity, and predictive values, key requirements for effective early warning systems^15,16^.

Here, we propose a two-stage co-learning architecture based on a stacking ensemble framework to address these limitations. Stacking extends conventional ensemble methods by introducing a supervised meta-classifier that learns how to optimally combine the outputs of multiple base models, rather than relying on fixed rules or weights^17^. In this study, binary outputs from multiple outbreak detection models are used as input features to train meta-classifiers, including logistic regression, extreme gradient boosting, and multilayer perceptron, to generate a unified early warning signal. This approach provides a flexible mechanism to reconcile conflicting alerts, reduces dependence on individual models, and improves detection consistency across heterogeneous epidemiological settings. The inclusion of both linear and non-linear models also enables evaluation of trade-offs between interpretability and predictive performance.

## Methods

### Data Sources

Weekly ILI encounter data were obtained from the Brazilian Health Information System for Primary Care (SIAPS, Ministry of Health, Brazil), covering all 5,570 Brazilian municipalities from November 2022 to July 2025. Encounters were identified using ILI-relevant codes from ICD-10 and ICPC-2^7,18^ (Table S1).

### Modelling Design

Our analysis follows five steps (Figure 1). Briefly, we calibrated seven ODMs on municipal ILI time series, generating weekly binary alarm signals. These alarms were then integrated through an unsupervised majority voting (MV) rule and three supervised meta-classifiers (logistic regression (LR), extreme gradient boosting (XGB), and multilayer perceptron (MLP)) trained to anticipate ILI surges defined by the Moving Epidemic Method (MEM). Performance was evaluated on a hold-out test period (2025) and through a simulation procedure with artificially inserted surges.

**Figure 1:**
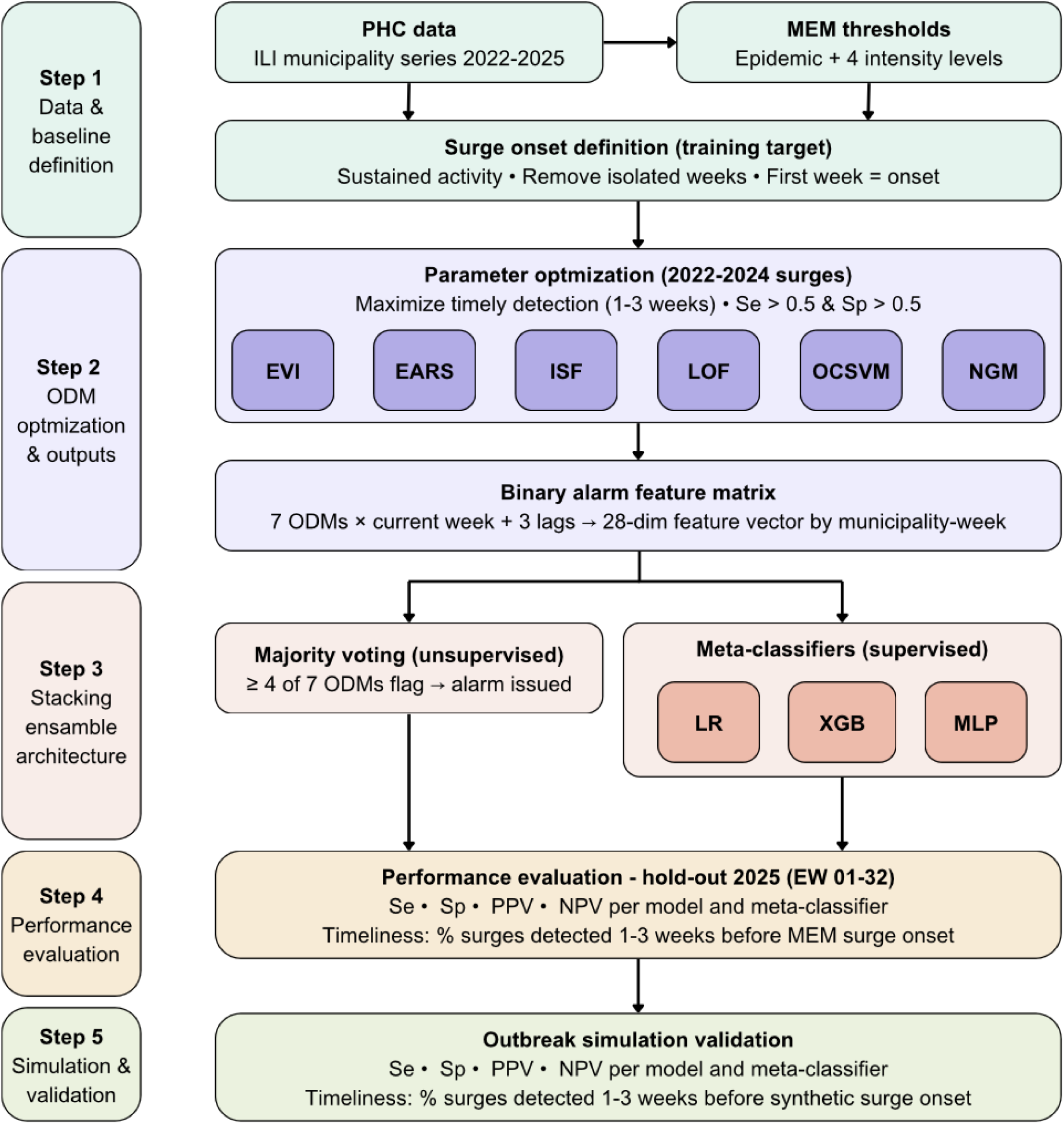
Two-stage stacking ensemble framework for influenza-like illness surge detection. The pipeline covers five steps, from data and baseline definition through model optimisation and stacking ensemble architecture to performance evaluation and simulation validation.

### Baseline ILI surges definition

We adopt the MEM to identify epidemic activity within ILI PHC time series^19^. The MEM uses historical data to estimate an epidemic threshold and four intensity levels (low, medium, high, and very high) based on mean values and confidence intervals, enabling us to flag both onset and severity characterisation of an outbreak. Thresholds were estimated from 2022–2024 data and applied prospectively to 2025. Isolated weeks above the epidemic threshold were excluded as they likely represent transient fluctuations. Consecutive exceedance weeks, defined as two or more weeks occurring consecutively or separated by a single non-exceedance week, were grouped into periods of sustained ILI activity, with the first week of each period defined as the surge onset^7^. Intensity classification details are provided in the Text S1.

### Outbreak Detection Models

We selected seven commonly used ODMs that represent a broad spectrum of detection principles: the Epidemic Volatility Index (EVI)^20^, the Early Aberration Reporting System (EARS) C2^21^, the Isolation Forest (ISF)^22^, the Local Outlier Factor (LOF)^23^, the One Class Support Vector Machine (OCSVM)^24^, the Copula-Based Outlier Detection (COPOD)^25^, and the Next Generation Method (NGM)^11^. Each model produces a weekly binary alarm indicating unusual ILI activity. Formulations, key parameters, warning definitions and assumptions are summarised in Table S2.

### ODM Model parametrization

Key parameters for each ODM were optimised using ILI surge events from 2022–2024, with 2025 reserved as an external benchmark. Parameter selection maximised timely detection, the proportion of surges preceded by an alarm one to three weeks in advance, subject to sensitivity (Se) and specificity (Sp) both exceeding 0·5, ensuring performance above chance while prioritising lead time. Se = TP/(TP + FN) and Sp = TN/(TN + FP), where TP (true positives) are alarms issued during or up to three weeks before increased ILI activity, TN (true negatives) are non-exceedance weeks without alarms, FN (false negatives) are exceedance weeks without alarms, and FP (false positives) are alarms during non-exceedance weeks.

After selecting the optimal parameters, the ODMs were re-evaluated over the full study period and their output served as inputs to the meta-classifiers. This approach avoids using the most recent data to calibrate the models and better reflects the constraints of real-time syndromic surveillance, where surges must be anticipated before they are observable in the surveillance data.

### Stacking Ensemble Framework

A stacking ensemble uses the outputs of multiple base models as features for a supervised second-stage meta-classifier, learning context-dependent combinations rather than fixed weights or simple aggregation rules^17^. For each municipality-week *i*, the feature vector comprises the current and three lagged binary alarms from each of the seven ODMs, yielding a 28-dimensional input. Each meta-classifier estimates *P*(*y_i_* = 1| *x_i_*), the probability of a surge at week *i*, and predictions are binarised at a 0·5 decision threshold. LR models a linear log-odds relationship between alarm patterns and surge probability, with L2 regularisation to prevent overfitting^26,27^. XGB builds an additive ensemble of decision trees, capturing nonlinear alarm interactions^28^. MLP learns complex input-output mappings through one hidden layer with nonlinear activation^29^. The MV rule issues an alarm when four or more ODMs flag the same week, providing an unsupervised baseline for comparison^11^. Full mathematical specifications are provided in the Text S2.

### Surge definition validation

MEM-derived surge definitions reflect historical baselines that may not capture all outbreak patterns. Since our modelling framework requires a target indicator, we complemented the hold-out evaluation with a simulation procedure. Following Noufaily *et al*.^4^, we generated 32 replicate series per municipality, each containing one to four artificial surges with random onset timing (weeks 1–32 of 2025), random duration (4–10 weeks), and outbreak size drawn from a Poisson distribution with mean kσ, where σ is the pre-2025 time series standard deviation and k is a scaling constant (1–10) controlling intensity. This design evaluates model sensitivity to surges of varying magnitude that may deviate from the MEM baseline^4,6,11^. Performance was assessed using Se, Sp, positive and negative predictive values (PPV and NPV, respectively)^6,7^.

## Results

### ILI encounters surges across Brazil

Between epidemiological week (EW) 42 of 2022 and EW 32 of 2025 (comprising 147 EWs), Brazil recorded a total of 1,257,818,754 PHC encounters. Of these, 66,818,181 were associated with ILI, representing 5·31% of all PHC encounters during the study period. At the city level, ILI-related encounters accounted for an average of 6·02% (95% CI: 5·93–6·11) of total encounters, corresponding to a mean of 11,994 (95% CI: 10,345–13,643) ILI cases out of an average of 225,780 (95% CI: 196,647–254,912) total encounters per city.

During the study period, we identified 5,365 cities with at least one ILI associated surge. Thus, after initial screening, 205 cities (3·9%) were excluded from subsequent ODMs and stacking ensemble analyses. From the valid cities, we identified a total of 137,474 city-weeks where ILI encounters exceeded the epidemic threshold. By aggregating consecutive epidemic periods, we identified 26,411 distinct surge onsets. Of these, 20,646 onsets (totaling 104,391 epidemic city-weeks) occurred prior to 2025, while 5,765 onsets (33,083 epidemic city-weeks) were recorded during 2025. Low- and medium-intensity surges accounted for 60·0% of the total onsets. The remaining surges were classified as high or very high intensity, as shown in Table 2 and Figure 2.

**Figure 2:**
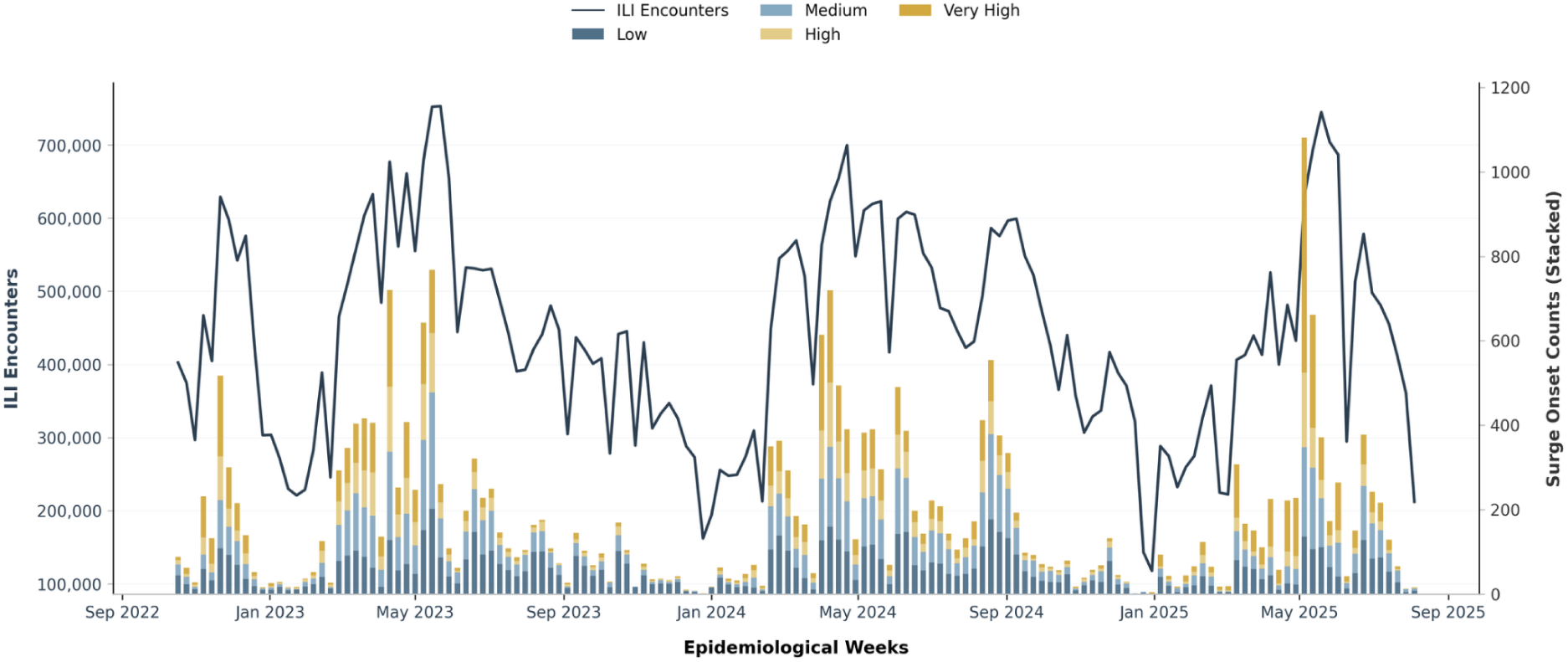
Temporal relationship between weekly ILI encounters and surge onsets in Brazil by intensity level (EW 42, 2022 – EW 32, 2025). Weekly ILI-related encounters counts are shown as the line curve (left y-axis). Stacked foreground bars quantify simultaneous surge onset counts (right y-axis), color-coded sequentially to reflect increasing severity profiles: low, medium, high, and very high intensity.

**Table 2:** Frequency and intensity distribution of ILI surge in Brazil by study period (EW 42/2022 to EW 32/2025).

|  | All period | Before 2025 | During 2025<br>(up to EW 32) |
| --- | --- | --- | --- |
| **Total surge onsets** | 26,411 | 20,646 | 5,765 |
| **Intensity** |  |  |  |
| Low | 8,244 (31·2%) | 6,825 (33·0%) | 1,422 (24·6%) |
| Medium | 7,605 (28·8%) | 6,106 (29·6%) | 1,498 (26·0%) |
| High | 4,041 (15·3%) | 3,308 (16·0%) | 732 (12·7%) |
| Very High | 6,521 (24·7%) | 4,407 (21·4%) | 2,113 (36·7%) |

Analysis of surges between EW 01 and EW 32 reveals a significant transition in intensity during 2025. While the relative frequency of low, medium, and high-intensity surges decreased significantly compared to 2023–2024 (p-value < 0·01), the proportion of very high intensity surges rose sharply to 36·7% in 2025 (p-value < 0·01), indicating a shift toward more severe surge events (<u>Table S3</u>).

### ODMs and meta-classifiers surge detection

After optimization of ODMs parameters (Table S3), an average of 30,603 (95% CI: 21,697–39,508) anomaly alerts across city-weeks were generated by the ODMs, ranging from 23,049 alerts for COPOD to 49,654 for EARS. Despite issuing a similar number of alerts overall, ODMs exhibited considerable heterogeneity in the anomalies they detected, with some methods identifying complementary patterns while others showed substantial agreement (Text S3).

In comparison, the meta-classifiers and the MV approach generated an average of 44,430 (95% CI: 26,426–62,435) anomaly alerts. Among them, the Multi-layer Perceptron (MLP) produced the highest number of alerts (52,264), followed by XGB (49,946) and LR (47,836), whereas MV generated the fewest (27,676).

### Timing Anticipation

The anticipation capacity of each model is illustrated in Figure 3A. Meta-classifiers demonstrated superior anticipatory capabilities compared to individual ODMs and achieved a substantial reduction in the proportion of missed surges, maintaining failure rates below 10%, whereas individual models like ISF and OCSVM missed over 30%.

**Figure 3:**
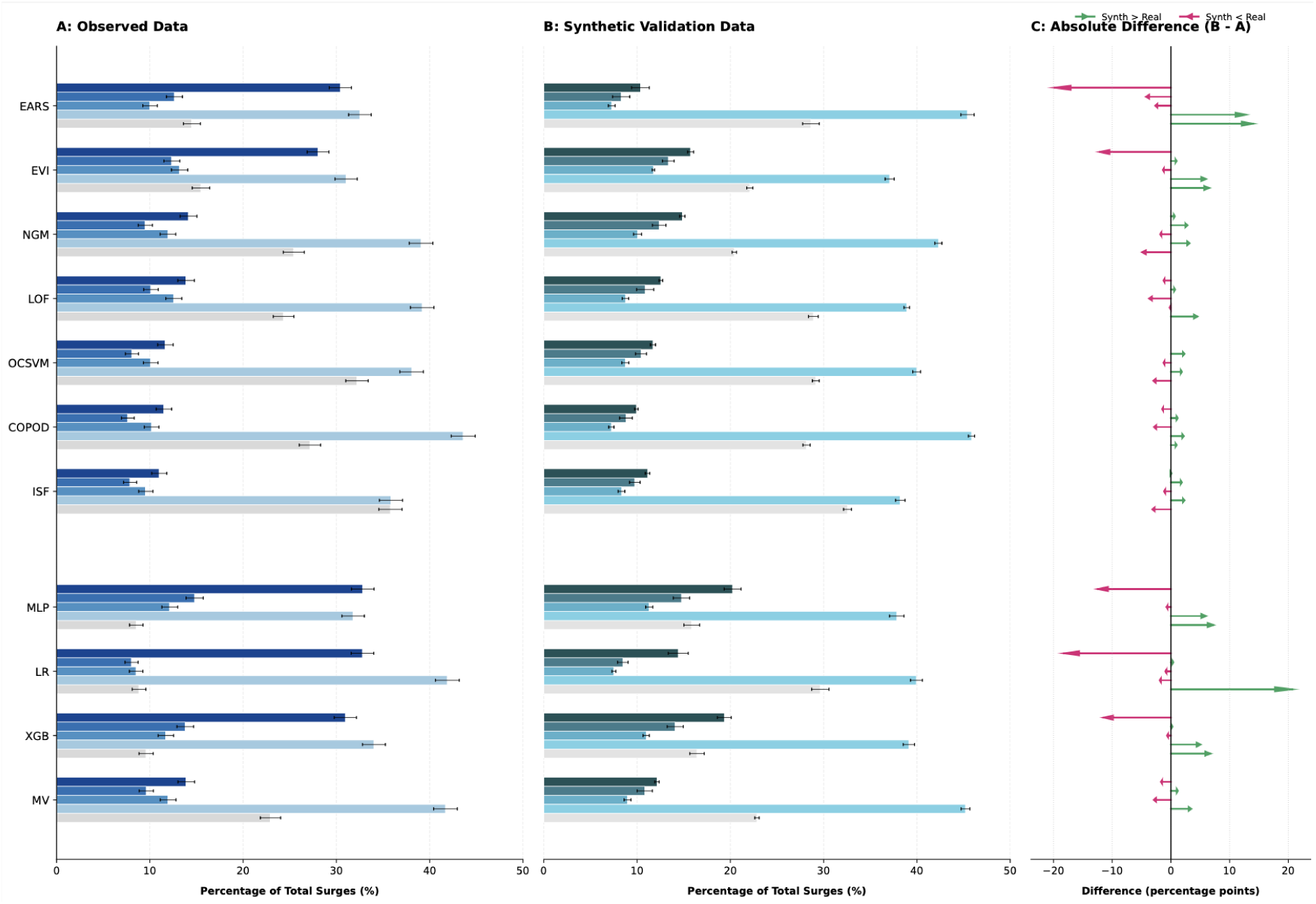
Timing anticipation of ODMs, meta-classifiers and MV. The bar charts show the percentage of surge onsets detected at different lead times (dark to light bars) and missed detections (light grey bars). Error bars indicate 95% confidence intervals. Panels show the anticipation timing of surge onsets in: (A) the original time series, using epidemic periods defined by MEM; (B) the simulated outbreak scenario, independent of the MEM threshold; and (C) the difference in detection percentages between the two scenarios (original series vs simulated synthetic surges), expressed in percentage points.

In the critical 3-week anticipation window, the LR and MLP anticipated 32·8% (95% CI: 31·6–34·0) of the total 5,765 surge onsets and XGB 30·9% (95% CI: 29·8–32·2). This represents an average increase of 15% for 3 weeks anticipation capacity with the meta-classifiers over individual models. In particular, while EARS also shows high 3-week anticipation (30·4%, 95% CI: 29·2–31·6), it suffers from a much higher missed rate (14·5%, 95% CI: 13·6–15·4) compared to the meta-classifiers.

In contrast, the MV technique performs much more like a traditional ODM than the other meta-classifiers. It has a high missed rate 22·9% (95% CI: 21·8–24·0) and lower anticipation performance (Figure 3A).

All models tend to miss more surges of low intensity and fewer surges of very high intensity. In particular, the relative distribution of detections across the timeline, occurring three, two, one weeks earlier, or in the same week, remains consistent with the general patterns observed for total surge counts (Figure S1), regardless of the intensity level.

ODMs missed an average of 32·4% (95% CI: 26·8%–38·1%) of low-intensity surges. Within this group, EVI achieved the highest sensitivity, missing the lowest percentage at 24·1% (95% CI: 23·2%–25·0%), while ISF missed the highest proportion (41·3%; 95% CI: 40·2%–42·4%).

The average percentage of missed low-intensity surges for ODMs was reduced by an average of 15·5% when using XGB, LR, and MLP classifiers. The meta-classifiers missed an average of 16·9% (95% CI: 10·8%–23·1%) of low-intensity surges. Among them, XGB missed the highest proportion (19·5%; 95% CI: 18·6%–20·3%), while MLP and LR missed 16·8% (95% CI: 16·0%–17·6%) and 14·5% (95% CI: 13·8%–15·3%), respectively. Finally, the MV missed 29·0% (95% CI: 28·0%–30·0%) of low-intensity surges.

In contrast, ODMs miss an average of 17·4% (95% CI: 12·0%–22·7%) of very high-intensity surges. Among these, EARS is the most effective at capturing these events, missing only 10·3% (95% CI: 9·6%–11·0%), while ISF shows the lowest sensitivity, with 26·6% of surges missed (95% CI: 25·6%–27·7%).

The XGB, LR, and MLP classifiers exhibit a substantially lower miss rate for very high-intensity surges, averaging 5·1% (95% CI: 4·6%–5·6) for MLP, 6·4% (95% CI: 5·8%–7·0%) for XGB and 6·8% (95% CI: 6·2%–7·5%) for LR. Finally, the MV missed 13·9% (95% CI: 13·1%–14·8%) of surges in this category.

### Average Model performance

Meta-classifiers demonstrated significantly superior sensitivity compared to ODMs. Meta-classifiers achieved a mean sensitivity of 91·0% (95% CI: 89·7–92·4%), whereas ODMs averaged 75·0% (95% CI: 67·7–82·3%). The MV model occupied an intermediate position with a sensitivity of 77·1% (95% CI: 76·0–78·2%) (Figure 4).

**Figure 4:**
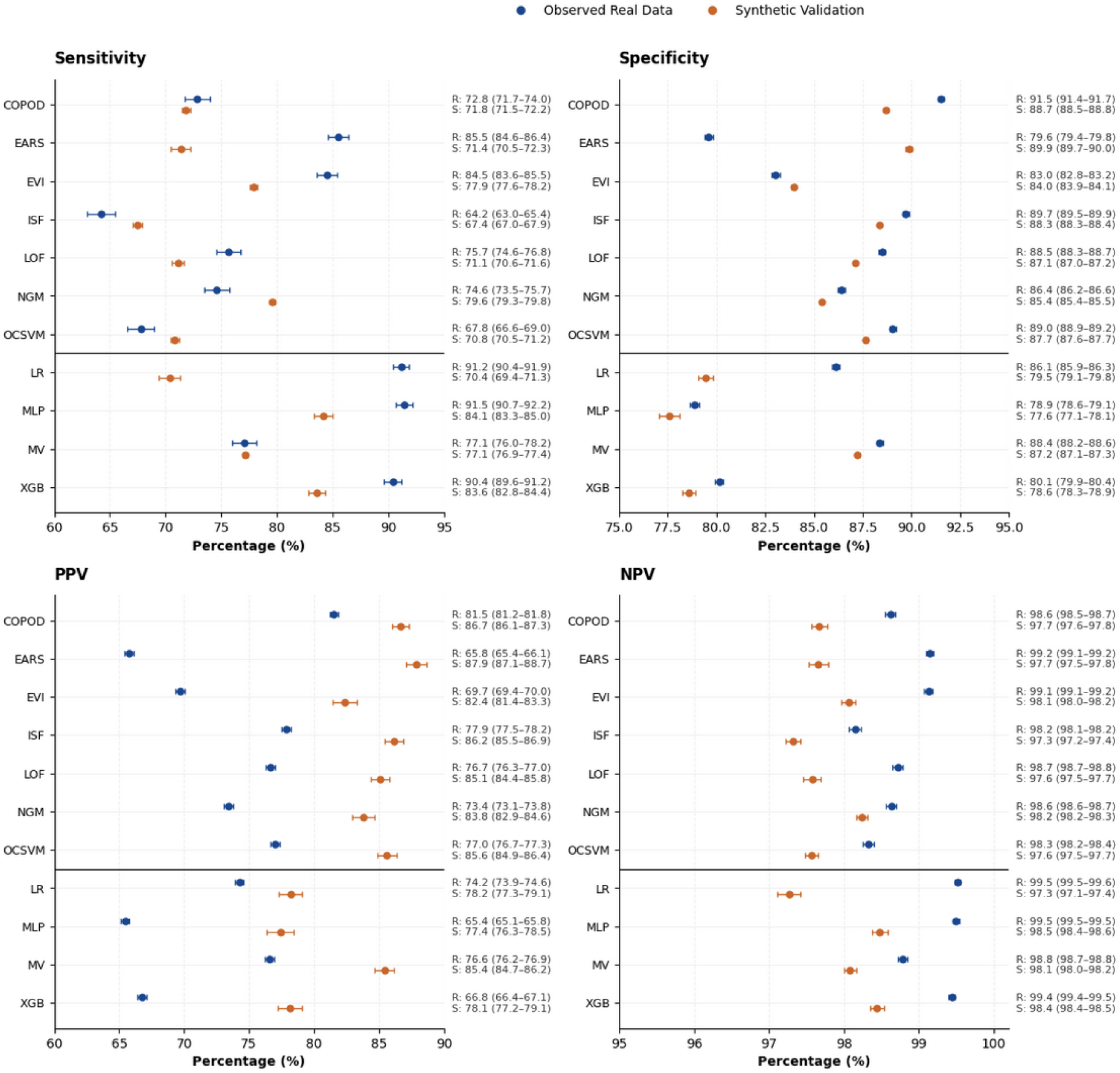
Validation performance of ODMs and meta-classifiers across observed and synthetic data frameworks. Panels display model performance across four diagnostic metrics: sensitivity (upper left), specificity (upper right), PPV (lower left), and NPV (lower right). Circular markers indicate mean estimates, with horizontal bars representing 95% CIs. Blue markers correspond to evaluations using observed data, whereas orange markers represent validation using synthetic datasets. Numerical estimates and corresponding 95% confidence intervals for both observed (O) and synthetic (S) frameworks are shown on the right side of each panel. The bold horizontal line separates individual ODMs (upper section) from ensemble meta-classifiers (lower section).

While meta-classifiers also provided a marginal improvement in NPV (99·5% vs. 98·7% for ODMs), this came at the cost of a slightly lower specificity and PPV. Compared with the meta-classifiers, ODMs showed higher average specificity (86·8% [95% CI: 83·0–90·7%] vs. 81·7% [72·1–91·4%]) and PPV (74·6% vs. 68·8%). The MV model showed specificity value of 88·4% (95% CI: 88·2–88·6%), PPV of 76·6% (95% CI: 76·2–76·9%), and NPV of 98·8% (95% CI: 98·7–98·9%)

### Surge definition validation

To conclude, we evaluated the impact of using MEM as a criterion for determining epidemic periods in ILI time series. During the 2025 period, our algorithm randomly inserted one or two synthetic surges into each replicate of the original time series across the 5,365 municipalities analyzed. This simulation produced a mean of 7,628 (95% CI: 7,396–7,858) surge onsets per replicate, compared to 5,765 onsets identified by MEM in the original series. Simulated outbreaks yield a mean of 59,806 (95% CI: 58,269–61,342) city-weeks classified as epidemic periods in the replicated series.

Relative to MEM epidemic thresholds estimated from the original series, synthetic surges were distributed across intensity categories as follows: 1,240 (95% CI: 1,213–1,267) low-intensity surges, 862 (95% CI: 840–883) medium-intensity surges, 371 (95% CI: 359–383) high-intensity surges, and 1,006 (95% CI: 960–1,052) very-high-intensity surges. Additionally, 4,149 (95% CI: 3,923–4,376) surges remained below the MEM epidemic threshold (sub-threshold surges). Consequently, only 45·6% of surge onsets, those exceeding the MEM threshold derived from the original data, were used to train the supervised meta-classifiers.

Accordingly, the number of anomalies issued in the synthetic series by individual ODMs ranged from a mean of 26,631 (95% CI: 26,494–26,769) for ISF to 36,751 (95% CI: 36,691–36,811) for EVI. Meta-classifiers generated a higher number of anomalies, with LR issuing a mean of 54,147 (95% CI: 53,859–54,436) anomalies, followed by MLP with 53,047 (95% CI: 52,302–53,791), XGB with 50,847 (95% CI: 50,512–51,183), and MV with the lowest mean of 29,944 (95% CI: 29,778–30,111) (Figure S2).

Despite the more challenging simulated scenario, meta-classifiers maintained improved performance compared with individual ODMs (Figure 3B). The proportion of surges detected three weeks in advance ranged from a mean of 12·3% (95% CI: 12·0–12·7) for ODMs to 18·0% (95% CI: 17·1–18·9) for meta-classifiers, whereas MV achieved 12·1% (95% CI: 11·9–12·4). Conversely, ODMs showed a mean missed-detection proportion of 27·1% (95% CI: 26·7–27·6), compared with 15·9% (95% CI: 15·0–16·7) for MLP, 16·4% (95% CI: 15·7–17·2) for XGB, 29·6% (95% CI: 28·7–30·6) for LR, and 22·9% (95% CI: 22·6–23·1) for MV.

Similar to before, all models tend to miss more sub-threshold intensity surges and fewer very high-intensity surges (Figure 5). In particular, between 40% and 60% of sub-threshold intensity surges are detected in the same week, with missing rates varying from 20% to 40% across all models. Notably, MLP and XGB meta-classifiers follow an expected trend of missing fewer surges at very high intensities, while ODMs, LR and MV retain higher than 10% missed rates for very high intensity surges.

**Figure 5:**
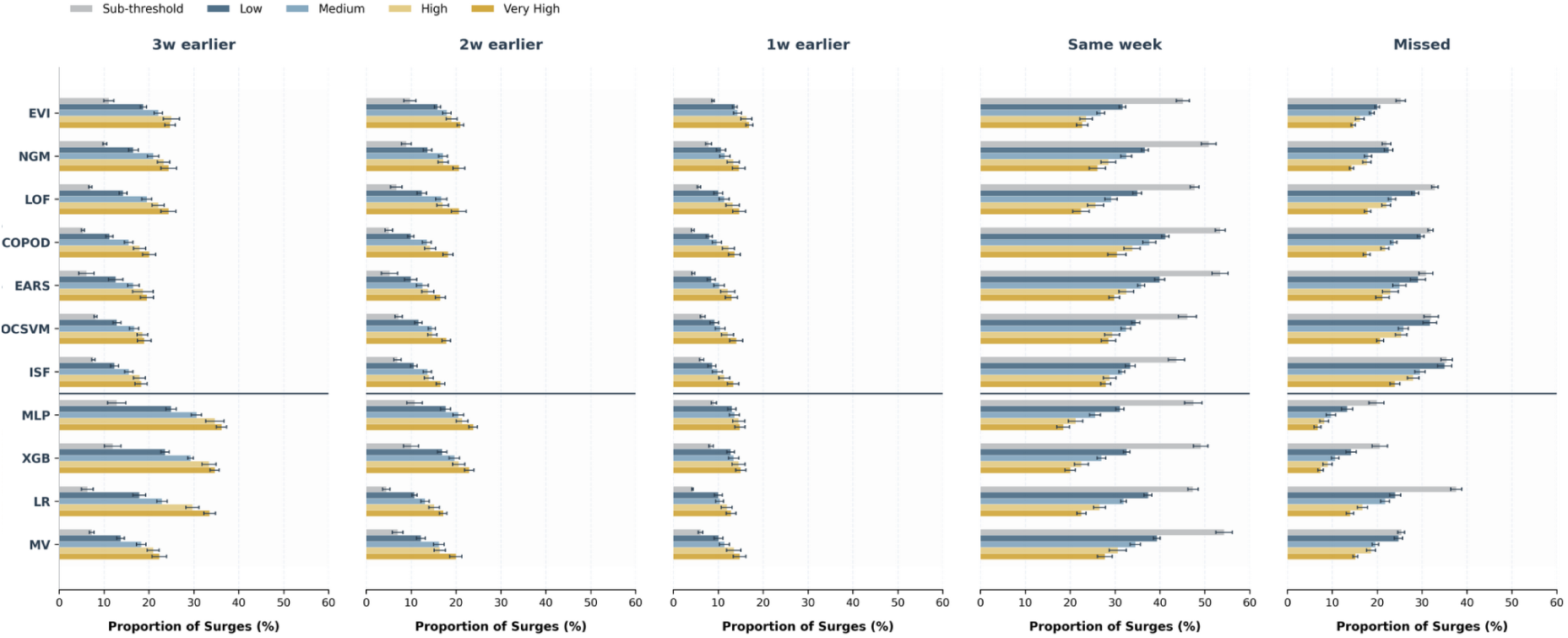
Comparative analysis of timeliness detection at different levels of surge intensity. The figure displays detection rates for ODMs (top) and Meta-classifiers (bottom) for total counts and across five intensity categories (Sub-threshold, Low, Medium, High and Very High). Bars indicate the percentage of total surges detected at specific lead times (3 weeks earlier to same week) or missed. Error bars represent 95% confidence intervals. Colors represent surge intensity.

Comparisons between observed and simulated analyses showed less than 10% variation in anticipation and missed-detection proportions for NGM, LOF, OCSVM, COPOD, ISF, and MV (Figure 3C). Notably, NGM demonstrated improved anticipation performance and lower missed-detection proportions compared with EARS. Similarly, MLP and XGB exhibited relatively stable performance across scenarios, with variations below 10% for most metrics, except for the proportion of surges detected three weeks in advance, which increased by 13·4% and 12·7%, respectively. In contrast, LR showed greater variability, with a 16·5% change in anticipation performance and a 20·1% change in missed-detection proportion between observed and synthetic scenarios.

Under the simulated scenario, performance metrics remained robust, with mean estimates and corresponding 95% confidence intervals consistently above 65% for sensitivity, 75% for specificity and PPV, and 97% for NPV.

## Discussion

A two-stage stacking ensemble framework, integrating multiple outbreak detection models, improved both the timeliness and predictive performance of ILI surge detection relative to any individual ODM, while preserving a strong balance across sensitivity, specificity, PPV, and NPV. The gains were consistent across surge intensity levels and were maintained under a simulated scenario with surges independent of the MEM baseline. This reinforces the reliability and applicability of this approach in early warning syndromic surveillance systems.

The need to move beyond single-model approaches has been recognised in the literature^4–7,15^. Bédubourg and Le Strat *et al.*^6^ assessed 21 ODMs on simulated surveillance data and concluded that no single algorithm achieved consistently adequate performance across all outbreak scenarios, highlighting the importance of methods that meet predefined minimum performance standards across varying conditions. Our results extend this finding: even when individual ODMs are parametrised to maximise timely detection against the same baseline, substantial variation persists in anticipation capacity, missed-detection rates, and sensitivity to surge intensity. These complementary characteristics are precisely what a stacking ensemble is designed to exploit. The MLP and XGB meta-classifiers improved timeliness and sensitivity (>80%) through adaptive weighting of individual model outputs while preserving specificity (>75%), PPV (>65%), and NPV (>98%). Importantly, this adaptive weighting was context-sensitive: EVI and NGM contributed more to low intensity surges and sub-threshold detection, while EARS weighted more heavily for high- and very-high-intensity surges, a pattern that would be invisible to any fixed aggregation rule.

This complementarity also extends to the temporal and spatial structure of alarms. EARS and EVI exhibited markedly lower correlation with the remaining ODM cluster, suggesting they may capture anomalies at different lead times or in municipalities where local data characteristics produce distinct signal dynamics. Whether this reflects genuine earlier detection in specific epidemiological contexts, or sensitivity to data artefacts, remains an open question that warrants investigation, particularly given the implications for how independent modelling approaches could be selectively weighted in geographically diverse surveillance systems^30^.

Simpler integration strategies performed less well. Majority voting behaved comparably to individual ODMs, as expected from a consensus-based rule that cannot exploit complementary signal timing across models. Logistic regression, while computationally simpler, showed the least stability across simulated scenarios, likely reflecting its assumption of linear separability between alarm-pattern combinations, an assumption that may not hold when surge baseline definitions shift. These findings suggest that supervised meta-learning (such as MLP and XGB), rather than aggregation alone, is necessary to realise the full benefit of model combination.

A recognised limitation of ODM evaluation is the absence of a universally accepted gold standard for surge definition^5–7^. Some studies use hospitalisation data, restricting evaluation to severe cases and missing the spectrum of mild cases that may still warrant public health attention^7,18^. Others use ensemble-labelling approaches or expert-derived definitions, which, particularly when developed with local health managers, can be highly appropriate for validating context-specific surveillance systems^13,31,32^. However, precisely because they are grounded in local knowledge and priorities, such definitions are difficult to standardise across regions, limiting their utility as a common reference for comparing surveillance performance in different settings.

We used the MEM, a widely adopted epidemiological method, to define epidemic thresholds and intensity levels from routine PHC data^19,33^. The method provides a replicable baseline that can be applied consistently across municipalities with different epidemiological profiles. Under the simulated scenario, where surges were generated independently of MEM and over half fell below the epidemic threshold, MLP and XGB maintained reliable and superior performance despite being trained on fewer than 50% of simulated events. These findings suggest that the framework is robust to alternative surge definitions, a property that becomes particularly valuable when combined with co-designed surveillance approaches. Studies such as Marcilio *et al.*^32^ have demonstrated how syndromic surveillance systems developed with local health managers can translate community-specific priorities into actionable surveillance targets. Our framework is compatible with this paradigm: the MEM epidemic threshold can be locally recalibrated to reflect what constitutes a surge of concern in a given setting, and the meta-classifiers can then learn to detect precisely those events. This creates a pathway toward early warning systems that are both methodologically rigorous and responsive to the epidemiological and operational realities of the communities they serve.

Several limitations warrant acknowledgement. The framework was evaluated on ILI in Brazilian municipalities; generalisability to other syndromes, data types, and health systems requires further study. Re-training the meta-classifiers requires labelled historical data and technical capacity that may not be available in all settings, and the operational trade-offs between model complexity and local implementation feasibility deserve prospective evaluation. Nevertheless, by establishing a shared reference framework, the stacking ensemble enables independent modelling teams to contribute separately to event detection within a unified analytical structure, an approach that could support distributed, multi-institutional surveillance networks and warrants further development.

In conclusion, integrating multiple modelling approaches through a stacking ensemble framework improves the timeliness and predictive performance of outbreak detection without requiring additional data infrastructure, offering a scalable methodological upgrade to existing syndromic surveillance pipelines.

## Data Availability

All data produced in the present work are contained in the manuscript

## Acknowledgements

This study is part of the Alert-Early System of Outbreaks with Pandemic Potential (ÆSOP, http://aesop.health), an initiative under development by Brazil’s Fundação Oswaldo Cruz (Fiocruz) and the Federal University of Rio de Janeiro with financial support from the Rockefeller Foundation’s Health Initiative (Grant 2023-PPI-007 awarded to MB-N). MB-N, PIPR and RFSA are CNPq Research Fellows. DGFB is a CNPq postdoctoral fellow (150420/2026-9). The excellent project management and administrative support from Fiotec (Foundation for Scientific and Technological Development in Health) is greatly appreciated. The National Institute of Science and Technology in Digital Health (DigiSaude-INCT) is supported by CNPq (408775/2024-6).

## Data sharing statement

Our agreement with the Brazilian Ministry of Health for accessing the referenced databases patently denies authorization of access to any third parties. All requests to access these databases must be addressed to the Brazilian Ministry of Health.

## Code availability

All generated data and codes to ensure reproducibility of our results are available in our GitHub repository https://github.com/cidacslab/aesop-detection-models.git.

## Contributions

Conceptualization: J.F.O., ALA, ERC, I.M, P.I.P.R., M.B.-N.; Methodology: J.F.O, ALA, ERC, DGFB, FMHSF, RSS, PTVF, MCSLC; Investigation: J.F.O., I.M., R.F.S.A., P.I.P.R. and M.B.-N.; Visualization: J.F.O. and RSS.; Funding acquisition: M.B.-N.; Project administration: I.M., P.I.P.R and M.B.-N.; Supervision: M.B.-N.; Writing—original draft: J.F.O., RSS and. M.B.-N; Writing—review and editing: all authors.

## References

1 Overview of Syndromic Surveillance What is Syndromic Surveillance? 2004; published online Sept 24. https://www.cdc.gov/mmwr/preview/mmwrhtml/su5301a3.htm (accessed Feb 12, 2026).

2 Babanejaddehaki G, An A, Papagelis M. Disease outbreak detection and forecasting: A review of methods and data sources. ACM Trans Comput Healthc 2025; 6: 1–40.

3 Ramos PIP, Marcilio I, Bento AI, et al. Combining digital and molecular approaches using health and alternate data sources in a next-generation surveillance system for anticipating outbreaks of pandemic potential. JMIR Public Health Surveill 2024; 10: e47673.

4 Noufaily A, Enki DG, Farrington P, Garthwaite P, Andrews N, Charlett A. An improved algorithm for outbreak detection in multiple surveillance systems. Stat Med 2013; 32: 1206–22.

5 Noufaily A, Morbey RA, Colón-González FJ, et al. Comparison of statistical algorithms for daily syndromic surveillance aberration detection. Bioinformatics 2019; 35: 3110–8.

6 Bédubourg G, Le Strat Y. Evaluation and comparison of statistical methods for early temporal detection of outbreaks: A simulation-based study. PLoS One 2017; 12: e0181227.

7 Oliveira JF, Cerqueira-Silva T, Brito PAN, et al. Anticipating influenza-like illness outbreaks via syndromic surveillance using over-the-counter drug sales and primary health care data. npj Digit Public Health 2026; 1: 10.

8 Xu L, Zhou C, Luo S, Chan DK, McLaws M-L, Liang W. Modernising infectious disease surveillance and an early-warning system: The need for China’s action. Lancet Reg Health West Pac 2022; 23: 100485.

9 Du L, Pang Y. A novel data-driven methodology for influenza outbreak detection and prediction. Sci Rep 2021; 11: 13275.

10 Wang P, Zhang W, Wang H, et al. Predicting the incidence of infectious diarrhea with symptom surveillance data using a stacking-based ensembled model. BMC Infect Dis 2024; 24: 265.

11 Borges DGF, Coutinho ER, Cerqueira-Silva T, et al. Combining machine learning and dynamic system techniques to early detection of respiratory outbreaks in routinely collected primary healthcare records. BMC Med Res Methodol 2025; 25: 99.

12 Carnevale RJ, Talbot TR, Schaffner W, Bloch KC, Daniels TL, Miller RA. Evaluating the utility of syndromic surveillance algorithms for screening to detect potentially clonal hospital infection outbreaks. J Am Med Inform Assoc 2011; 18: 466–72.

13 Saab A, Dabboussi AH, Abi Khalil C, Rahme J, Salem Sokhn E, El Morr C. Practical approach for evaluating machine learning anomaly detection algorithms for epidemic Early Warning Systems. Stud Health Technol Inform 2025; 327: 1205–9.

14 Florentino PTV, Bertoldo Junior J, Barbosa GCG, et al. Impact of Primary Health Care data quality on infectious disease surveillance in Brazil: Case study. JMIR Public Health Surveill 2025; 11: e67050.

15 Meckawy R, Stuckler D, Mehta A, Al-Ahdal T, Doebbeling BN. Effectiveness of early warning systems in the detection of infectious diseases outbreaks: a systematic review. BMC Public Health 2022; 22: 2216.

16 Steele L, Orefuwa E, Dickmann P. Drivers of earlier infectious disease outbreak detection: a systematic literature review. Int J Infect Dis 2016; 53: 15–20.

17 Sagi O, Rokach L. Ensemble learning: A survey. Wiley Interdiscip Rev Data Min Knowl Discov 2018; 8: e1249.

18 Cerqueira-Silva T, Oliveira JF, Oliveira V de A, et al. Early warning system using primary health care data in the post-COVID-19 pandemic era: Brazil nationwide case-study. Cad Saude Publica 2024; 40: e00010024.

19 Vega T, Lozano JE, Meerhoff T, et al. Influenza surveillance in Europe: establishing epidemic thresholds by the moving epidemic method. Influenza Other Respi Viruses 2013; 7: 546–58.

20 Kostoulas P, Meletis E, Pateras K, et al. The epidemic volatility index, a novel early warning tool for identifying new waves in an epidemic. Sci Rep 2021; 11: 23775.

21 Hutwagner L, Thompson W, Seeman GM, Treadwell T. The bioterrorism preparedness and response Early Aberration Reporting System (EARS). J Urban Health 2003; 80: i89–96.

22 Liu FT, Ting KM, Zhou Z-H. Isolation Forest. In: 2008 Eighth IEEE International Conference on Data Mining. IEEE, 2008: 413–22.

23 Breunig MM, Kriegel H-P, Ng RT, Sander J. Lof: identifying density-based local outliers. In: Proceedings of the 2000 ACM SIGMOD international conference on Management of data. 2000: 93–104.

24 Schölkopf B, Platt JC, Shawe-Taylor J, Smola AJ, Williamson RC. Estimating the support of a high-dimensional distribution. Neural Comput 2001; 13: 1443–71.

25 Li Z, Zhao Y, Botta N, Ionescu C, Hu X. COPOD: Copula-Based Outlier Detection. In: 2020 IEEE International Conference on Data Mining (ICDM). IEEE, 2020. DOI:10.1109/icdm50108.2020.00135.

26 Yu H-F, Huang F-L, Lin C-J. Dual coordinate descent methods for logistic regression and maximum entropy models. Mach Learn 2011; 85: 41–75.

27 LogisticRegression. scikit-learn. https://scikit-learn.org/stable/modules/generated/sklearn.linear_model.LogisticRegression.html (accessed March 3, 2026).

28 Introduction to Boosted Trees — xgboost 3.2.0 documentation. https://xgboost.readthedocs.io/en/stable/tutorials/model.html (accessed March 3, 2026).

29 Coutinho ER, Silva RM, Delgado ARS. Utilização de Técnicas de Inteligência Computacional na Predição de Dados Meteorológicos. Rev Bras Meteorol 2016; 31: 24–36.

30 de Oliveira JF. Mathematical modelling as a tool for precision health equity. Nat Med 2024; 30: 1802–3.

31 Hicketier A, Bach M, Oedi P, Ullrich A, Abbood A. Ensemble-labeling of infectious diseases time series to evaluate early warning systems. 2024. DOI:10.2139/ssrn.4940608.

32 Marcilio I, Ramos PIP, Florentino PVT, et al. Implementation of AESOP early-warning system for respiratory disease: a pilot and validation study using routinely collected data in Amazonas, Brazil. Lancet Reg Health Am 2026; 60: 101516.

33 Teeluck M, Samura A. Assessing the appropriateness of the Moving Epidemic Method and WHO Average Curve Method for the syndromic surveillance of acute respiratory infection in Mauritius. PLoS One 2021; 16: e0252703.

