## Supplementary Materials for "Enhancing Early Warning Outbreak Detection Using Multi Model Stacking Ensemble"

### The PDF file includes:

### Supplementary Figures 1 and 2

Fig. S1: Comparative analysis of timeliness detection at different levels of surge intensity.

Fig. S2: Mean number of warnings triggered by each evaluated model on replicated city-level time series.

### Supplementary Table 1 to 3

Table S1: ICD-10 and ICPC-2 codes used to define influenza-like illness encounters.

Table S2: Overview of ODMs used to evaluate anomalous weeks of ILI associated PHC encounters in each city level timeseries.

Table S3: Frequency and intensity distribution of ILI surge onsets in Brazil restricted to the period from EW 01 to EW 32.

### Supplementary Text 1 and 4

Text S1: Outbreak Intensity classification

Text S2: Mathematical formulations of the meta-classifier models

Text S3:Hierarchical clustering and correlation analysis of ODMs

Text S4: Supplementary references

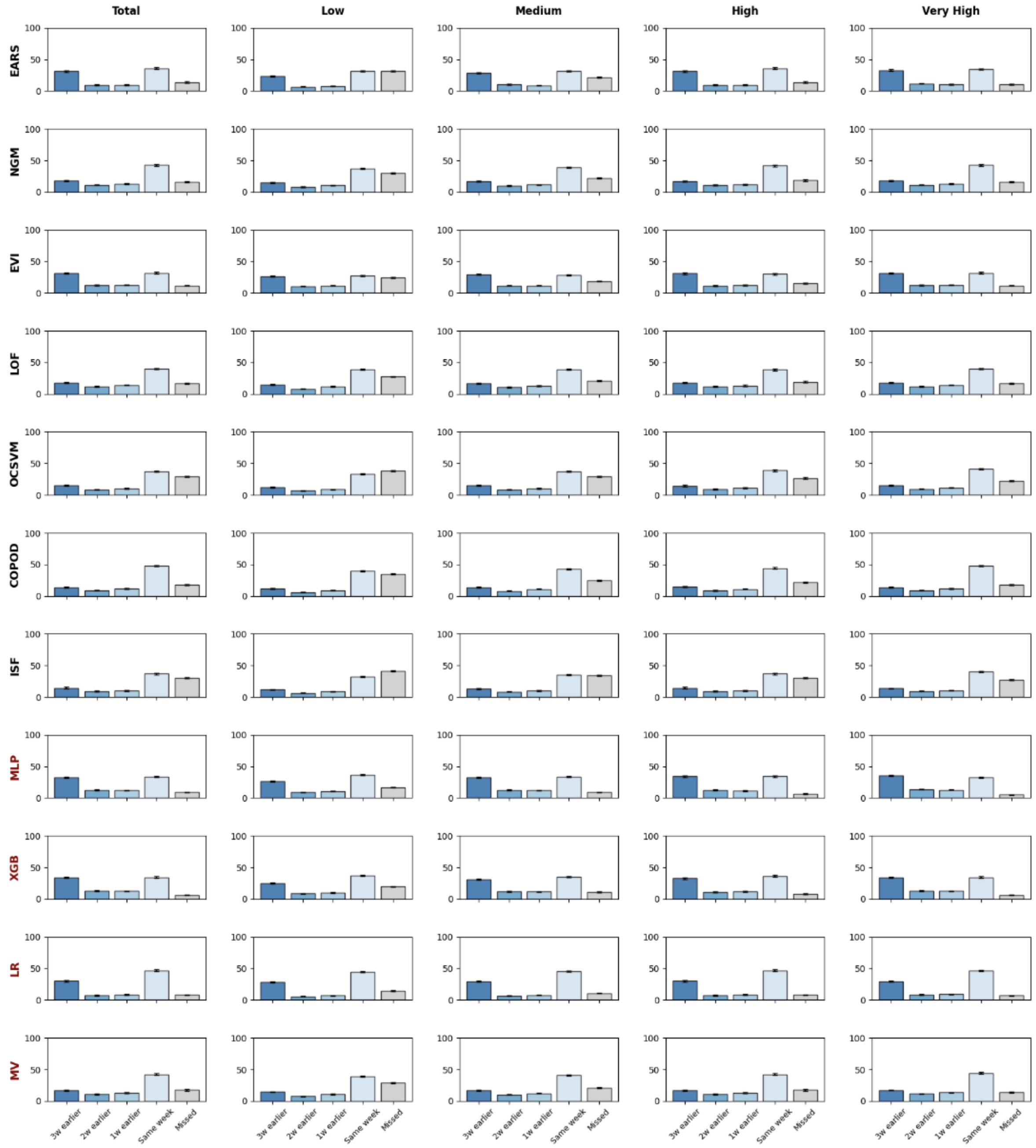

**Figure S1: Comparative analysis of timeliness detection at different levels of surge intensity.** The figure displays detection rates for ODMs (top) and Meta-classifiers and MV (bottom) for total counts and across four intensity categories (Low, Medium, High and Very High). Bars indicate the percentage of total surges detected at specific lead times (3 weeks earlier to same week) or missed. Error bars represent 95% confidence intervals. Colors represent lead-time detection, from dark blue (3 weeks earlier) to grey (missed).

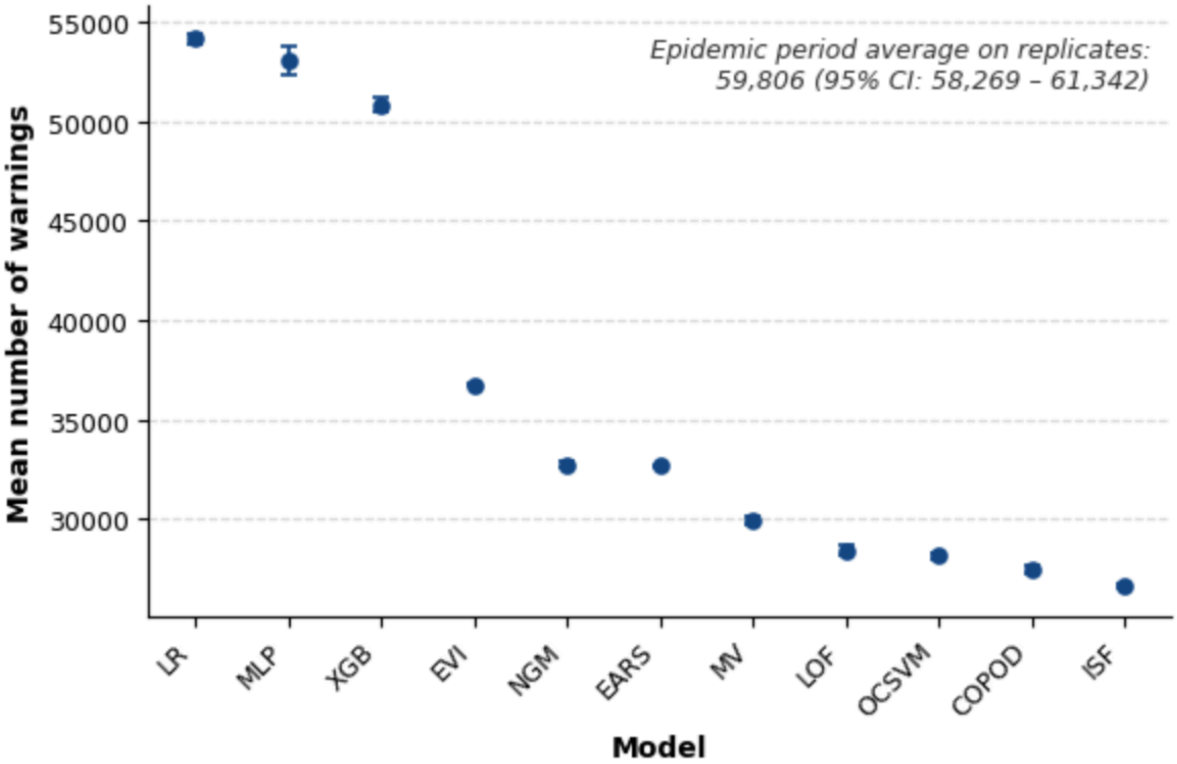

**Figure S2: Mean number of warnings triggered by each evaluated model on replicated city-level time series.** Error bars denote 95% confidence intervals. The top-right inset indicates the epidemic period average across replicates (59,806; 95% CI: 58,269–61,342).

**Table S1**: **ICD-10 and ICPC-2 codes used to define influenza-like illness encounters.** Codes for PHC encounters are extracted from the SIAPS classified using the International Classification of Diseases (ICD-10) and the International Classification of Primary Care (ICPC-2).

| **Type** | **Code** | **Description** |
|---|---|---|
| ICPC-2 | A03 | Fever |
| ICPC-2 | R01 | Pain respiratory system |
| ICPC-2 | R02 | Shortness of breath/dyspnoea |
| ICPC-2 | R03 | Wheezing |
| ICPC-2 | R04 | Breathing problem, other |
| ICPC-2 | R05 | Cough |
| ICPC-2 | R07 | Sneezing / nasal congestion |
| ICPC-2 | R08 | Nose symptom / complaint other |
| ICPC-2 | R21 | Sinus symptom / complaint |
| ICPC-2 | R23 | Voice symptom / complaint |
| ICPC-2 | R25 | Sputum / phlegm abnormal |
| ICPC-2 | R29 | Respiratory symptom/complaint other |
| ICPC-2 | R71 | Whooping cough |
| ICPC-2 | R74 | Upper respiratory infection acute |
| ICPC-2 | R75 | Sinusitis acute/chronic |
| ICPC-2 | R76 | Tonsillitis acute |
| ICPC-2 | R77 | Laryngitis/tracheitis acute |
| ICPC-2 | R78 | Acute bronchitis/bronchiolitis |
| ICPC-2 | R80 | Influenza |
| ICPC-2 | R81 | Pneumonia |
| ICPC-2 | R83 | Respiratory infection other |
| ICPC-2 | R99 | Respiratory disease other |
| ICD-10 | J00 | Acute nasopharyngitis |
| ICD-10 | J01 | Acute sinusitis |
| ICD-10 | J02 | Acute pharyngitis |

| | | |
|---|---|---|
| ICD-10 | J03 | Acute tonsillitis |
| ICD-10 | J04 | Acute laryngitis and tracheitis |
| ICD-10 | J06 | Acute upper respiratory infections of multiple and unspecified sites |
| ICD-10 | J09 | Influenza due to identified zoonotic or pandemic influenza virus |
| ICD-10 | J10 | Influenza due to identified seasonal influenza virus |
| ICD-10 | J11 | Influenza, virus not identified |
| ICD-10 | J12 | Viral pneumonia, not elsewhere classified |
| ICD-10 | J13 | Pneumonia due to Streptococcus pneumoniae |
| ICD-10 | J14 | Pneumonia due to Haemophilus influenzae |
| ICD-10 | J15 | Bacterial pneumonia, not elsewhere classified |
| ICD-10 | J16 | Pneumonia due to other infectious organisms, not elsewhere classified |
| ICD-10 | J17 | Pneumonia in diseases classified elsewhere |
| ICD-10 | J18 | Pneumonia, organism unspecified |
| ICD-10 | J20 | Acute bronchitis |
| ICD-10 | J21 | Acute bronchiolitis |
| ICD-10 | J22 | Unspecified acute lower respiratory infection |
| ICD-10 | J80 | Adult respiratory distress syndrome |
| ICD-10 | R05 | Cough |
| ICD-10 | R06 | Abnormalities of breathing |
| ICD-10 | R07 | Pain in throat and chest |
| ICD-10 | R43 | Disturbances of smell and taste |
| ICD-10 | R50 | Fever of other and unknown origin |
| ICD-10 | U07 | Emergency use of U07 |
| ICD-10 | B34 | Viral infection of unspecified site |
| ICD-10 | B97 | Viral agents as the cause of diseases classified to other chapters |

**Table S2:** Overview of Outbreak Detection Models used to evaluate anomalous weeks of associated ILI PHC encounters in each city level timeseries.

| **Model** | **Summary** | **Key formulation and parameters** | **Warning definition** | **Assumptions** |
|---|---|---|---|---|
| Epidemic Volatility Index (EVI)[1] | The EVI model is based on the standard deviation growth rate calculated over consecutive rolling windows of a time series. This defines the $EVI_{t-1,t}$ index, which captures sudden increases in epidemic volatility by measuring the change in variability between two adjacent windows ($t-1$ and $t$). | $m$: size of the rolling window (number of consecutive observations), with $0 < m < n$, where $n$ is the length of the time series. This results in $n - m + 1$ overlapping rolling windows.<br><br>$c$: volatility threshold ($c \in [0, 1]$) used to identify above-average epidemic volatility, defined as $EVI_{t-1,t} \geq c$. | A warning is issued at week t with observation $y_t$, when both conditions are satisfied:<br>(i) $EVI_{t-1,t} \geq c$; and<br>(ii) the newly observed value $y_t$ exceeds the average number of reported cases over the last seven recent weeks. | Optimal values for $m$ and $c$ are selected using ILI surge events from 2022–2024. |
| Early Aberration Reporting System (EARS-C2)[2] | The EARS method is available in four versions (EARS C-1 to 3 and EARS-NB). Here EARS-C2 is chosen based on Cerqueira-Silva et al.[3]. The EARS-C2 monitors time series by computing a moving sample average ($\mu_2$) and a sample standard deviation ($\sigma_2^2$). | $\mu_2$: moving sample average given by<br>$$\mu_2(t) = \frac{1}{7}\sum_{i=t-3}^{t-9} y_i,$$<br>where $y_i$ is an observation count at time $t = i$.<br><br>$\sigma_2^2$: sample standard deviation<br>$$\sigma_2^2(t) = \frac{1}{6}\sum_{i=t-3}^{t-9} (y_i - \mu_2(t))^2$$<br>$C_2$: the residual given by<br>$$C_2(t) = \frac{y_t - \mu_2(t)}{\sigma_2(t)}$$<br><br>α: is a value in the unit interval [0,1] used to establish the posterior bound for $C_2$. | It is assumed that in the absence of an outbreak, the residual $C_2$ follows a standard normal distribution. Thus, a warning is issued at week $t$ with observation $y_t$, if:<br>$$C_2 \geq z_{1-\alpha}$$<br>where $z_{1-\alpha}$ is the $(1-\alpha)^{th}$ - quantile of the standard normal distribution. | The EARS-C2 threshold used for the definition of warnings may be adjusted to achieve a required model's accuracy. |

| | | | | |
|---|---|---|---|---|
| Isolation Forest (ISF)[4] | The ISF method is based on the concept of isolation, which means the ability to separate an observation from the rest of the data. The algorithm recursively partitions the dataset using randomly selected features and random split values. Each recursive partition can be represented as a binary decision tree (iTree), where the number of splits needed to isolate an observation, $y = y_t$, corresponds to the path length ($h(y)$) of that observation within the tree.<br><br>By constructing a collection of such trees (the "forest"), the model computes the average path length for each observation. | $n_{estimators}$: Defines the number of trees in the forest. A higher value tends to increase the robustness and stability of predictions, but also increases computational cost.<br><br>*contamination*: Estimates the proportion of anomalies in the dataset, influencing the determination of the score threshold. It is a crucial parameter for adjusting the model's sensitivity. | The ISF model first converts path length into scores to issue a warning, as<br><br>$s(y, n) = 2^{-\frac{E(h(y))}{\mu(n)}}$,<br><br>with $\mu(n) = 2H(n - 1) - (2(n - 1)/n$ being the average path length of the iTree, n is the time series length, $H(i) \approx ln(i) + Euler's\ constant$ is a harmonic number and $E(h(y))$ is the average of $h(y)$ in the forest.<br><br>Higher scores of $s$ correspond to more anomalous observations. In particular, observations with shorter average path lengths throughout the forest, that is, points that usually require fewer splits to isolate, are anomalies. | 1) There is no need to optimize the model's parameters.<br><br>2) Contamination values less than 0.5 are a good way to approximate the true anomaly proportion, as stated in Liu et al.[4]. |
| Local Outlier Factor (LOF)[5] | The LOF is a density-based anomaly detection method that quantifies how isolated a data point is with respect to the local neighborhood surrounding it. LOF evaluates whether a point lies in a region of substantially lower density than that of its nearest neighbors.<br><br>Let $N_k(a)$ be the set of k-nearest neighbors of a point $a$. The LOF construction starts with the k-distance of a point, which defines the neighborhood radius used to characterize local density. Based on this quantity, the reachability distance between points $a$ and $b$ is defined as | $n_{neighbors}$: Defines the number of neighbors $N_k$ used to estimate the local density around each observation. Larger values produce smoother and more stable density estimates, but may reduce sensitivity to small localized deviations. Smaller values, in contrast, make the method more responsive to local irregularities, although also more sensitive to noise. Therefore, this parameter directly controls the scale at which "locality" is defined.<br><br>*contamination*: As with ISF, this parameter specifies the expected | The LOF score of a data point $a$ is calculated from the ratio between the local reachability density of its neighbors and its own local reachability density. Formally, it is defined as<br><br>$LOF(a) = \frac{\sum_{b \in N_k(a)} \frac{LRD_k(b)}{LRD_k(a)}}{\lvert N_k(a) \rvert}$<br><br>This factor measures how much less dense the region around a is when compared with the regions occupied by its neighbors. If LOF $(a) \approx 1$, then point a has a local density comparable to that of its neighborhood and is | 1) There is no need to optimize the model's parameters.<br><br>2) The LOF method relies on the assumption that normal observations are embedded in neighborhoods with relatively homogeneous local density, whereas anomalous observations occur in regions whose density is substantially lower than that of nearby points. |

| | | | | |
|---|---|---|---|---|
| | $reach - dist_k(a, b) = max\{k - distance(b), d(a,b)\}$<br><br>where $k - distance(b)$ is the notation representing the shortest distance from $b$ to its k-th neighbour, $d(a, b)$ is the distance between $a$ and $b$. This definition prevents very small neighbor distances from producing unstable density estimates.<br><br>The local reachability density (LRD) of point a is then given by the inverse of the average reachability distance from a to its neighbors:<br><br>$LRD_k(a) = \left( \frac{\sum_{b \in N_k(a)} reach - dist_k(a,b)}{\lvert N_k(a) \rvert} \right)^{-1}$ | proportion of anomalies in the dataset and is used to determine the decision threshold that separates normal observations from anomalous ones. Thus, it does not change the LOF score itself, but affects the binary classification derived from that score. | therefore consistent with the surrounding data structure. In contrast, values LOF $(a) > 1$ indicates that the point lies in a region of lower density than its neighbors, suggesting anomalous behavior. The larger the LOF value, the stronger the evidence that the point is a local outlier. Thus, an alert is triggered when the LOF score exceeds a limit greater than 1, that is, when the point is classified as anomalous according to the contamination-adjusted decision limit. | 3) LOF assumes that the selected number of neighbors k is adequate to capture the relevant local structure of the data. If k is too small, the score may become unstable and overly sensitive to noise; if k is too large, genuinely local anomalies may be smoothed out and remain undetected. |
| One-Class Support Vector Machine (OCSVM)[6] | OCSVM is an unsupervised anomaly detection method designed to learn the boundary of a reference class composed predominantly of normal observations. OCSVM estimates a decision function that encloses most data points in the training set within a compact region of the feature space, while points lying outside this region are regarded as anomalies.<br><br>To achieve this, the original input data are mapped into a high-dimensional feature space through a kernel function Φ. In that space, the algorithm searches for a hyperplane that maximally separates the mapped data from the origin. This construction allows OCSVM to | ν*:* Controls the trade-off between the flexibility of the decision boundary and the tolerance to outliers. In the OCSVM formulation, ν, in the interval $(0, 1]$ acts as an upper bound on the fraction of training observations allowed to fall outside the learned boundary and as a lower bound on the fraction of support vectors. Larger values of ν generally produce a more restrictive boundary and increase the sensitivity to anomalous observations.<br><br>*kernel (RBF):* Specifies the kernel function used to project the data into the feature space. The radial basis | To separate the target data from the origin in the feature space, OCSVM solves an optimization problem that seeks a hyperplane with maximum margin while allowing some observations to violate the boundary through slack variables. The margin is defined as<br><br>$M = \frac{b}{\lvert\lvert w \rvert\rvert}$<br><br>where $w$ is the normal vector to the hyperplane and $b$ is the bias term. The primal optimization problem is given by | 1) The OCSVM assumes that the training data are composed predominantly of normal observations and that these observations occupy a relatively coherent region in the feature space. In other words, the method relies on the idea that normal data can be enclosed by a boundary, whereas anomalies are expected to fall outside this learned support.<br><br>2) Another important assumption is that the |

| | | | | |
|---|---|---|---|---|
| | characterize the support of the distribution of normal data, especially when the underlying structure is nonlinear. | function (RBF) kernel is a common choice because it can capture nonlinear patterns and complex decision boundaries, which are often required in anomaly detection problems.<br><br>γ: Determines the width of the RBF kernel and, consequently, the locality of the model. Small values of γ generate smoother and more global decision boundaries, whereas larger values make the boundary more sensitive to local variations in the data. | $min_{(w, b, \xi)}(\frac{1}{2}\|\|w\|\|^2 + \frac{1}{\nu n}\sum_{i=1}^{n}\xi_i - b)$<br><br>subject to<br><br>$(w \cdot \Phi(x_i)) \geq b - \xi_i, i = 1, \cdots,$ [illegible]<br><br>with $\xi_i \geq 0$ for all $i = 1, \cdots, n$.<br><br>In this formulation, $n$ denotes the number of training observations, $\xi_i$ are slack variables that allow boundary violations, and ν controls the trade-off between maximizing the distance of the hyperplane from the origin and tolerating points outside the learned support.<br><br>After training, the anomaly score is determined from the decision function<br><br>$f(x) = (w \cdot \Phi(x) - b)$.<br><br>Equivalently, in kernel form, the decision rule can be expressed in terms of the support vectors and kernel evaluations. A point x is considered normal when it lies inside the learned decision region, i.e., when $f(x) \geq 0$, and anomalous when it falls outside this region, i.e., when $f(x) < 0$. Therefore, a warning is triggered whenever the decision function assumes a negative value, indicating that the observation is incompatible | kernel function is capable of representing the structure of the normal class adequately. In this sense, model performance depends strongly on the choice of kernel and its parameters, particularly γ in the RBF case. If γ is too small, the decision boundary may become excessively smooth and fail to capture important structures; if it is too large, the model may overfit local fluctuations and become overly sensitive to noise.<br><br>3) There is no need to optimize the model's parameters. |

| | | | | |
|---|---|---|---|---|
| | | | with the support estimated from the normal data. | |
| Copula-Based Outlier Detection (COPOD)[7] | COPOD is an unsupervised anomaly detection method based on the empirical representation of data distribution through copulas.<br><br>The method is grounded in copula theory, which allows the dependency structure between variables to be separated from their marginal distributions. In practice, each variable is transformed into its empirical cumulative distribution function (CDF), mapping the original data onto a normalized probability space. For a given observation, COPOD assesses whether its coordinates lie in the extreme lower or upper tails of the empirical distribution. Observations that simultaneously exhibit rare or extreme positions in one or more dimensions receive higher anomaly scores and are interpreted as potential outliers. | $contamination$: Specifies the expected proportion of anomalies in the dataset and is used to define the decision threshold for labeling obser-vations as anomalous. As in other unsupervised detectors, this parameter does not alter the raw anomaly score itself, but determines the cutoff used for binary classification. | Let $X = (X_1, \cdots, X_p)$ be a $p$-dimensional random vector. For each variable $X_j$, the empirical cumulative distribution function is used to evaluate the relative position of an observation $x_j$ within its marginal distribution. The lower-tail probability is given by<br><br>$$U_j(x_j) = F_j(x_j),$$<br><br>where $F_j(\cdot)$ denotes the empirical cumulative distribution function of the j-th variable.<br><br>Similarly, the upper-tail probability can be represented by<br><br>$$\overline{U}_j(x_j) = 1 - F_j(x_j).$$<br><br>To quantify how extreme the observation is in each dimension, COPOD uses tail-based scores derived from these probabilities. A common representation is<br><br>$s_j(x_j) = -log(min\{F_j(x_j), 1 - F$ [illegible]<br><br>so that values lying near either tail of the marginal distribution produce larger scores. The overall anomaly score of an observation $x$ is then obtained by aggregating the marginal tail scores | 1) COPOD assumes that anomalous observations are characterized by unusually extreme positions in one or more variables, particularly when these extremes occur in regions of low joint probability under the empirical dependence structure of the data. It is essentially parameter-light, since it does not require tuning hyperparameters such as the number of neighbors, tree depth, or kernel width. Its main mechanism relies on estimating the empirical marginal distributions of the variables and combining their tail probabilities to quantify abnormality.<br><br>2) COPOD assumes that anomalies are relatively rare compared with the bulk of normal observations. In practical implementations, this assumption is reflected by the contamination parameter, which specifies the expected proportion of anomalous observations and is used to define the decision threshold. If the |

| | | | | |
|---|---|---|---|---|
| | | | across dimensions, typically through a maximum or cumulative combination: $$S(x) = \sum_{j=1}^{p} s_j(x_j)$$ In all cases, higher values of $S(x)$ indicate that the observation occupies a low-probability region of the joint distribution and is therefore more likely to be anomalous. A warning is triggered when the aggregated COPOD score exceeds a prede-fined threshold or when the observation is classified as anomalous according to the contamination-adjusted cutoff. | dataset contains a large proportion of abnormal points, the empirical distributions may themselves become distorted, reducing the contrast between normal and anomalous patterns and potentially impairing detection performance. 3) There is no need to optimize the model's parameters. |
| Next Generation Method (NGM)[8] | The NGM is an epidemiological model that takes into account the time of infection based on the Next Generation Method (NGM), a mathematical approach used to estimate the time-dependent reproduction number ($R_t$), an important metric for quantifying the transmission dynamics of infectious diseases over time. The current implementation uses the SEIR compartmental model and defines a generation interval distribution that describes the time required for an infected individual to cause a new infection. We estimate the discretized value of $R_t$, for $t = i$ by: | δ: Defines the threshold bounding $R_t$ for identifying a significant change in the behavior of the time series of observed counts.. γ: Recovery rate. | A warning is issued at week t with observation $y_t$, when both conditions are satisfied: (i) $R_t > \delta$ ; and (ii) the newly observed value $y_t$ exceeds the average number of reported cases over the last five recent weeks. | 1) The value for δ for a reproduction number greater than classically used (threshold = 1) is assumed for syndromic data. This threshold denotes the sensitivity and specificity, respectively, indicating the model's accuracy in detecting these increases in PHC encounters. 2) Optimal values for δ and γ are selected using ILI surge events from 2022–2024. |

| | |
|---|---|
| | $$R_i = \frac{y_i}{\sum_{j=1}^{n} g_i y_{(i-j)}}$$ where $y_i$ is the observed counts at week $t = i$ and $g_i$ is a generation interval distribution function. We assume a priori that the dynamics of the disease to be detected early are unknown, so the function $g_i = g(i)$ reduces to: $$g(i) = \gamma^2 i e^{-\gamma i}$$ |
| All methods are applied to a smoothed version of the time series (moving average). | |

**Table S3:** Frequency and intensity distribution of ILI surge onsets in Brazil restricted to the period  from EW 01 to EW 32 of each year.

| | **Total** | **2023** | **2024** | **2025** |
|---|---|---|---|---|
| **Total** | 20,543 | 7,403 | 7,375 | 5,765 |
| Low | 5,795 (28.2%) | 2,138 (28.9%) | 2,235 (30.3%) | 1,422 (24.7%) |
| Medium | 5,997 (29.2%) | 2,340 ( 31.6%) | 2,159 (29.3%) | 1,498 (26.0%) |
| High | 3,290 (16.0%) | 1337 (18.1%) | 1221 (16.6%) | 732 (12.7%) |
| Very High | 5,461 (26.6%) | 1588 (21.5%) | 1760 (23.9%) | 2,113 (36.7%) |

**Table S4: Key parameters for early anomaly detection models optimized/established for ILI encounter time series.** Parameter configurations for maximizing lead-time (anticipation) contingent on sensitivity and specificity exceeding 0.5.

| Model | Key optimised parameters |
|---|---|
| Epidemic Volatility Index (EVI) | $m$: 6.7 (95% CI: 5.58 – 6.83)<br><br>$c$: 0.58 (95% CI: 0.56 – 0.60) |
| Early Aberration Reporting System (EARS-C2) | α: 0,15<br><br>b = 10<br><br>offset = 3 |
| Isolation Forest (ISF) | $n_{estimators}$: 400<br><br>$contamination$: 0.5 |
| Local Outlier Factor (LOF) | $n_{neighbors}$: 400<br><br>$contamination$: 0.5 |
| One-Class Support Vector Machine (OCSVM) | ν*:* 0.8<br>*kernel: Radial Basis Function*<br>γ *:* 0.02 |
| Copula-Based Outlier Detection (COPOD) | $contamination$: 0.5 |
| Next Generation Method (NGM) | δ: 1.2<br><br>γ: 0.5 |

### Supplementary Text 1

Outbreak Intensity classification

The MEM uses historical surveillance data to calculate an epidemic threshold and four intensity levels (low, medium, high, and very high). Thresholds were established using data from 2022 to 2024 and applied prospectively to classify ILI activity in 2025. MEM intensity thresholds classify surges as: low intensity if the weekly number of ILI encounters exceeded the epidemic threshold but stayed below the low threshold; medium intensity if counts fell between the low and medium thresholds; high intensity if counts fell between the medium and high thresholds; and very high intensity if counts exceeded the high threshold.

MEM thresholds served as the benchmarks for defining activity levels across the entire study period. Specifically, a week was classified as having increased activity when the observed encounters exceeded the epidemic threshold; in 2025, these exceedances were interpreted as epidemic periods relative to the historical baseline.

### Supplementary Text 2

Mathematical formulations of the meta-classifier models

A stacking ensemble is a meta-learning technique in which a supervised second level model (meta-classifier) uses the outputs (features) of multiple base models as inputs. The goal is to learn context-dependent combinations of ODMs outputs, rather than applying fixed weights or voting rules[9]. Considering the type of ODM outputs, we implemented three supervised meta-classifiers to combine model anomalies: the Logistic Regression, XGB, and the MLP. In addition, we also assessed the majority voting approach to compare it with the meta-classifiers performances.

To provide a mathematical description for each classifier, consider the following notation: let $j \in \{1, ..., 8\}$ index the ODMs, $i \in \{1, ..., n\}$ index time points (weeks), so that $x_{ij} \in \{0, 1\}$ is the binary alarm generated by the $j$th ODM at week $i$, and $y_i \in \{0, 1\}$ is our target variable, i.e., the reference surge indicator (baseline surge definition).

To include the early anticipation capability of each classifier, we also added another temporal feature, namely the time-lags up to three weeks as $x_{ij}^{k} = x_{i-k,j}$, for $k \in \{1, ..., 3\}$. In this way, the vector of features, for each week $i$, becomes:

$$x_i = (x_{i1}^0, x_{i1}^1, x_{i1}^2, x_{i1}^3, \ldots, x_{i8}^0, x_{i8}^1, x_{i8}^2, x_{i8}^3)^T, \quad \text{for } x_{ij}^s \in \{0, 1\}$$

Therefore, each classifier uses the training data $\{x_i\}_{i=4}^{n}$ to predict $\{y_i\}_{i=4}^{n}$ and estimates the conditional probability:

$$\hat{y}_i = P(y_i = 1 | x_i)$$

which represents the probability of a surge in a week $i$, based on current and previous three weeks alarm patterns.

To translate the continuous probabilistic output of the meta-classifiers into an actionable binary warning, we applied a decision threshold (0.5) chosen to balance early detection capabilities against the risk of excessive warning.

In the sequence, we present a brief characterization of the three used supervised meta-classifiers.

Logistic Regression (LR)

It assumes a linear relationship between the log-odds of the outcome and the input features. The model is defined as:

$$P(y_i = 1|x_i) = \sigma(\beta_0 + \sum_{j=1}^{8}\sum_{k=0}^{3}\beta_{jk}x_{ij}^k)$$

where $\sigma = \frac{1}{1+e^{-z}}$ is the logistic function, $\beta_0$ is the intercept and the regression coefficients $\beta_{jk}$ measures the immediate and delayed predictive effects. In other terms, the regression coefficients explicitly represent the weighting of each ODM[10].

The model parameters are estimated with the L2 regularization[11]. Thus the objective function become:

$$\mathcal{L}(\theta) = -\frac{1}{n}\sum_{i=1}^{n}[y_i ln(\hat{y}_i) + (1-y_i)ln(1-\hat{y}_i)] + \frac{\lambda}{2}\theta^T\theta$$

where $\theta = [\beta_0, \beta_{jk}]$ is the vector of model parameters, for $k \in \{1, 2, 3\}$ and $j \in \{1, \cdots, 8\}$, $\hat{y}_i$ is the predicted probability of an event at time $i$, and $\lambda > 0$ controls regularization strength, which helps us to avoid overfitting.

Extreme Gradient Boosting (XGB)

XGB is a gradient boosting method that builds an additive ensemble of decision trees and can capture nonlinear interactions between alarms. The model is defined as:

$$y_i = \sigma(\sum_{s=1}^{S} f_s(x_i))$$

where $f_s$ are regression trees, $S$ is the number of trees, and $\sigma$ is the logistic function. Each tree partitions the feature space (combinations of alarm patterns) into regions and assigns a score to each region. The final prediction is the sum of tree outputs, transformed into a probability.

The model parameters are estimated by minimizing a regularized objective function:

$$\mathcal{L} = \sum_{i=1}^{n} l(y_i, \hat{y}_i^S) + \sum_{i=1}^{n}\Omega(f_s)$$

where $l(y_i, \hat{y}_i^S)$ is the training loss function, $S$ is the number of trees and $\Omega$ is a regularization term that penalizes tree complexity[12].

Multi-Layer Perceptron (MLP)

A MLP is a fully connected neural network composed of an input layer, one or more hidden layers, and an output layer. Each input neuron represents a feature, and every neuron in one layer connects to all neurons in the next. Hidden layers transform the input data through weighted connections and nonlinear activation functions, enabling the model to learn complex relationships between inputs and outputs. The output layer produces the final prediction, with the number of neurons corresponding to the number of target variables[13].

With one hidden layer, the model can be written as:

$$h_r = g(\omega_{r0} + \sum_{p=1}^{32}\omega_{rp}x_{ip})$$

being the hidden layer, and the output layer:

$$P(y_i = 1|x_i) = \sigma(\alpha_0 + \sum_{r=1}^{R} \alpha_r h_r)$$

where $g$ is an activation function, $R$ is the number of hidden neurons, σ is the logistic function.

When the network produces output, we calculate the loss function:

$$\mathcal{L} = -\frac{1}{n}\sum_{i=1}^{n}[y_i ln(\hat{y_i}) + (1 - y_i) ln(1 - \hat{y_i})]$$

The goal of MLP is then to minimize the loss function, by adjusting the network's weight and bias through backpropagation. We used the gradient descent method for this process.

Majority Voting (MV)

Finally, we also consider the MV method. In this approach, the final warning is generated according to what most models suggest. A warning is issued each week when 4 or more models have issued a warning.

Finally, epidemic surges are rare events compared to periods of baseline incidence, resulting in imbalanced datasets. To prevent the meta-classifiers from developing a bias towards the majority class (non-surge weeks), distinct balancing strategies were embedded into the training architectures. For the MLP, we applied the Synthetic Minority Over-sampling Technique (SMOTE) within the training folds via a pipeline structure, preventing data leakage into the validation and testing sets. For LR, class imbalance was addressed using the balanced class weight mode. For the XGB, class imbalance was addressed natively through Cost-Sensitive Learning. We applied a scaling weight to the positive class proportional to the negative-to-positive instance ratio observed in the training data (scale_pos_weight). Furthermore, to prevent the XGB algorithm from assigning excessive importance to the minority class and generating unstable probability estimates, we constrained the tree weight updates using a maximum delta step (max_delta_step).

### Supplementary Text 3

Hierarchical clustering and correlation analysis of ODMs

By examining the dynamics of ODMs models, we evaluated the degree of time-spatial overlap in anomalies across the different methods. Figure 1 below reveals a clear divergence between two groups of ODMs. The first group, comprising EARS and EVI, significantly deviates from the others. This suggests that EARS and EVI may be anticipating or lagging behind the anomalies identified by the other models, or perhaps capturing anomalies in different contexts driven by local data characteristics. Notably, EARS and EVI occupy isolated significant branches in the dendrogram, indicating they capture distinct patterns even from each other. EVI emerges as the most independent model, exhibiting the lowest mean correlation (0·37) with all other ODMs.

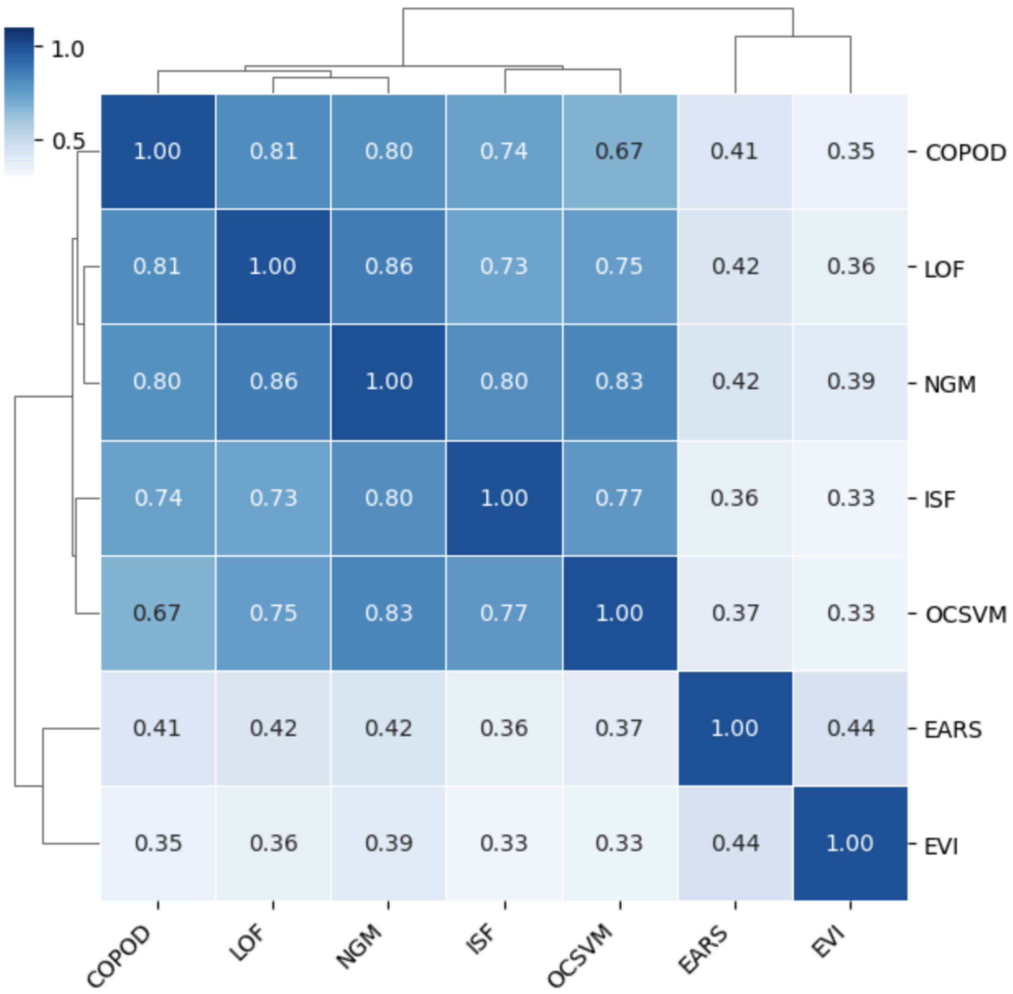

**Figure 1: Hierarchical clustering and correlation analysis of ODMs.** The heatmap displays the pairwise correlation coefficients between model outputs, with the dendrogram illustrating the emergence of how models are significantly differing.

Conversely, the second cluster of models (COPOD, LOF, NGM, ISF, and OCSVM) displays strong internal correlation, ranging from 0·68 to 0·88. This suggests high synchronicity in both time and space, implying that an anomaly detected by one model in this group is likely to be mirrored by the others within the same week and city. In particular, NGM and LOF are the most correlated models (0·86).

### Supplementary references

1 Kostoulas P, Meletis E, Pateras K, *et al.* The epidemic volatility index, a novel early warning tool for identifying new waves in an epidemic. *Sci Rep* 2021; **11**: 23775.

2 Hutwagner L, Thompson W, Seeman GM, Treadwell T. The bioterrorism preparedness and response Early Aberration Reporting System (EARS). *J Urban Health* 2003; **80**: i89–96.

3 Cerqueira-Silva T, Oliveira JF, Oliveira V de A, *et al.* Early warning system using primary health care data in the post-COVID-19 pandemic era: Brazil nationwide case-study. *Cad Saude Publica* 2024; **40**: e00010024.

4 Liu FT, Ting KM, Zhou Z-H. Isolation Forest. In: 2008 Eighth IEEE International Conference on Data

Mining. IEEE, 2008: 413–22.

5 Breunig MM, Kriegel H-P, Ng RT, Sander J. Lof: identifying density-based local outliers. In: Proceedings of the 2000 ACM SIGMOD international conference on Management of data. 2000: 93–104.

6 Schölkopf B, Platt JC, Shawe-Taylor J, Smola AJ, Williamson RC. Estimating the support of a high-dimensional distribution. *Neural Comput* 2001; **13**: 1443–71.

7 Li Z, Zhao Y, Botta N, Ionescu C, Hu X. COPOD: Copula-Based Outlier Detection. In: 2020 IEEE International Conference on Data Mining (ICDM). IEEE, 2020. DOI:10.1109/icdm50108.2020.00135.

8 Borges DGF, Coutinho ER, Cerqueira-Silva T, *et al.* Combining machine learning and dynamic system techniques to early detection of respiratory outbreaks in routinely collected primary healthcare records. *BMC Med Res Methodol* 2025; **25**: 99.

9 Sagi O, Rokach L. Ensemble learning: A survey. *Wiley Interdiscip Rev Data Min Knowl Discov* 2018; **8**: e1249.

10 Yu H-F, Huang F-L, Lin C-J. Dual coordinate descent methods for logistic regression and maximum entropy models. *Mach Learn* 2011; **85**: 41–75.

11 LogisticRegression. scikit-learn. https://scikit-learn.org/stable/modules/generated/sklearn.linear_model.LogisticRegression.html (accessed March 3, 2026).

12 Introduction to Boosted Trees — xgboost 3.2.0 documentation. https://xgboost.readthedocs.io/en/stable/tutorials/model.html (accessed March 3, 2026).

13 Coutinho ER, Silva RM, Delgado ARS. Utilização de Técnicas de Inteligência Computacional na Predição de Dados Meteorológicos. *Rev Bras Meteorol* 2016; **31**: 24–36.
